# Changes in sleep duration during the long-lasting COVID-19 pandemic: individual and regional disparities

**DOI:** 10.1101/2023.10.25.23297531

**Authors:** Jieun Min, Jieun Oh, Whanhee Lee

## Abstract

The adequate quality and quantity of sleep are related to maintaining the immune system and mental well-being; therefore, it is necessary to evaluate sleep duration during COVID-19. This study aimed to investigate the changes in sleep duration during the long-lasting COVID-19 period (2020 and 2021) in South Korea, and to examine the individual and regional disparities. The study population comprised 1,143,460 adults aged ≥19 years who participated in the 2017– 2021 Korea Community Health Survey excluding those who did not respond to the daily sleep duration questionnaire. For statistical analysis, we first conducted a multiple regression model for 229 districts to estimate the district-specific changes in sleep duration. We then applied a meta-analysis to pool the 229 estimates and a meta-regression to examine the association between changes in sleep duration and regional characteristics. The sleep duration increased by 9.66 (95% CI: 8.53, 10.80) min in 2020 and 3.66 (95% CI: 2.09, 5.22) min in 2021 compared to the pre-pandemic period (2017–2019). The increase was more prominent in males, younger adults, employed individuals, and those with a high socioeconomic status compared to the general population. Communities with a higher proportion of apartments, lower normalized difference vegetation index in summer, and lower practice rate of moderate exercise were associated with a higher increase in sleep duration during the pandemic. The sleep duration increased during the COVID-19 pandemic, and the increase decreased as the COVID-19 lasted longer. The findings of our study highlight that preventive measures to manage sleep health during a pandemic should be framed in consideration of individual and regional characteristics.

## Introduction

The Coronavirus disease 2019 (COVID-19) pandemic has posed an unprecedented threat to public health. To prevent the spread of the virus, lockdowns, social distancing, and quarantines have been implemented [1]. These intervention measures resulted in changes in people’s lifestyles by reducing social interaction and closing schools or workplaces, which may lead to symptoms of psychological distress [2, 3].

In the context of the COVID-19 pandemic, the importance of sleep in preventing risks to physical and mental health is widely being acknowledged. Adequate sleep helps maintain the immune system and reduces the risk of infectious diseases [4, 5]. In contrast, too little or too much sleep may lead to hypertension, metabolic and cardiovascular diseases, and increase the risk of death [6–9]. Sleep problems, including hypersomnia and sleep deprivation, are associated with psychiatric symptoms, such as depression and mood disorders, and can lower emotional regulation and cognitive processing capacity [10–14]. Therefore, it is necessary to evaluate sleep during the COVID-19 period to respond to physical and mental health-related problems.

Changes in sleep duration during the period of social distancing and stay-at-home orders have been reported in previous studies [15–21]. Sleep patterns changed in the United States and 16 European countries during the early stages of COVID-19 (Jan–Apr of 2020) [16]. People in five major metropolitan areas (London, Los Angeles, New York City, Seoul, and Stockholm) increased their average sleep duration in the months after the COVID-19 pandemic onset (Jan–Apr of 2020), compared to the same months in the prior year (Jan–Apr of 2019) [20]. Studies have found that the proportion of individuals who sleep more than the recommended duration of 7 hours has increased [18, 21].

In addition, sleep duration during the COVID-19 pandemic may be associated with individual and regional characteristics. Previous studies in the early period of the COVID-19 pandemic found different patterns by sex and age; however, the results were inconsistent [15, 17]. Heterogeneous associations between COVID-19 and sleep duration were also observed by socioeconomic status and residential areas; individuals with high education levels or living in urban and suburban areas were more likely to sleep more during the pandemic, although the impacts of occupational status were mixed [15].

Nevertheless, most previous studies regarding the relationship between COVID-19 and sleep outcomes were conducted in the early stages of the COVID-19 pandemic (i.e., 2020) and did not consider the nationwide population [15, 18, 20]. Moreover, although sleep duration and its changes can be heterogeneous based on various individual and regional characteristics, they have not been mainly investigated. Accordingly, this study aimed to investigate the changes in sleep duration during the long-lasting COVID-19 period (2020 and 2021) for the nationwide population in South Korea and to examine the related individual and regional disparities.

## Methods

### Study design and population

This was a cross-sectional study on changes in sleep duration during the COVID-19 pandemic (2020–2021) compared to the previous three years (2017–2019). The study population included the subjects who participated in the 2017–2021 Korea Community Health Survey (KCHS), which is an annual nationwide community-level health survey conducted by the Korea Centers for Disease Control and Prevention since 2008. The KCHS is a self-reported survey conducted on adults aged ≥19 years residing in all of the 229 districts in South Korea. The study participants were selected each year through probability proportionate and systematic sampling [22]. Among the all 1,144,331 survey participants of the 2017-2021 KCHS, this study examined 1,143,460 subjects, excluding those who did not respond to the daily sleep duration questionnaire.

### Sleep duration

In the KCHS, the daily mean sleep duration was measured in hours based on the following open-ended question in 2017–2019: “How many hours do you usually sleep a day?”. In 2020– 2021, the daily mean sleep duration was separately responded to for the weekdays and weekends; therefore, we calculated the weighted average of the daily mean sleep duration using the two questionnaires for the daily mean sleep duration including both the weekdays and weekends.

### Potential confounders

To consider the potential confounding effect, the following individual-level confounders were obtained from the KCHS: age group (19-39 y, 40-64 y, or 65+ y), sex (male or female), subjective health status (bad, normal, or good), smoking status (never, past-smoker, or current-smoker), drinking status (never, past-drinker, or current-smoker), household income (< 3 million won/month or ≥ 3 million won/month), education level (less than college or college or higher), marital status (married, [divorced, widowed, or separated], or never-married), economic activity status (no or yes), single-person household (no or yes), hypertension history (no or yes), and diabetes history (no or yes). We included participants who did not answer the questionnaire for confounders to consider their characteristics.

### District-level indicators

To estimate the associations between the changes in sleep duration and community characteristics, we used the following six district-level indicators during the study period: proportion of apartments among residences, proportion of mental counseling due to stress, local tax per person, normalized difference vegetation index (NDVI) in the summer, practice rate of moderate exercise, and number of bars per 1,000 people. The proportion of apartments among residences could represent the urbanization status because apartments are considered luxury residency type and proportion of apartments tend to be higher in city than in rural in South Korea. Proportion of mental counseling due to stress can partly show the mental well-being of each district and local tax per person is indicator of economic status. We collected NDVI, practice rate of moderate exercise, and number of bars because residential greenness, physical activity, and alcohol drinking is known to be associated with sleep quality [23–25]. Data regarding the local tax was obtained from the Korea Statistics Office, and the NDVI was obtained from the Moderate Resolution Imaging Spectroradiometer (MODIS). The remaining indicators were collected from the database of community health outcomes and determinants distributed by the Korea Centers for Disease Control and Prevention. All the indicators were averaged across the study period and included in the meta-regression model.

### Statistical analyses

The analysis was based on a two-stage analysis to estimate the changes in sleep duration during (2020 and 2021) and before (2017–2019) the COVID-19 pandemic and to determine whether these changes are associated with the community characteristics. All statistical analyses were conducted using the R statistical software (version 4.1.0).

In the first stage, district-specific time trend-adjusted changes in sleep duration comparing the period of the pandemic (2020 and 2021) to the timeframe before the pandemic (2017–2019) were calculated by fitting multiple regression models. A categorical variable indicating the pre-outbreak period (2017–2019), early stage (2020), and long-lasting period (2021) of the COVID-19 pandemic was included in the model to quantify district-specific changes in sleep duration during and before the pandemic. The year variable was linearly adjusted to control the temporal trends. The complex sampling survey structure was considered and all confounders were adjusted for in the first stage.

In the second stage, we conducted a random-intercept meta-analysis to pool the 229 district estimates to calculate the overall change in sleep duration during the pandemic in South Korea. To examine whether the changes in sleep duration are associated with community characteristics, we performed a meta-regression analysis with a random intercept using six district-level indicators. The longitude and latitude of each district were adjusted to alleviate unmeasured spatial heterogeneity. The estimated associations in meta-regression were expressed as an increase in the sleep duration for an interquartile range (IQR) increase of the district-level indicators.

### Sub-population analysis

To examine the disproportionate changes in sleep duration by individual characteristics, we conducted a sub-population analysis. We repeated the aforementioned two-stage analysis based on sex, age (19–39 y, 40–64 y, and 65+ y), economic activity status (no and yes), household income (<3 million won/month and ≥3 million won/month), and educational level (less than college and college or higher).

### Ethics statement

Informed consent was not required because publicly available and de-identified KCHS data was used.

## Results

With a total of 1,143,460 study participants, the average daily sleep duration was 6.61 hours a day before the outbreak of the COVID-19 pandemic (2017–2019) and it increased during the pandemic to 6.82 hours/day and 6.72 hours/day in 2020 and 2021, respectively (Table 1). Considering 229 districts, the median value of the proportion of apartments among the residences was 51.88%, demonstrating a relatively high variance across the districts (Table 2).

**Table 1.**
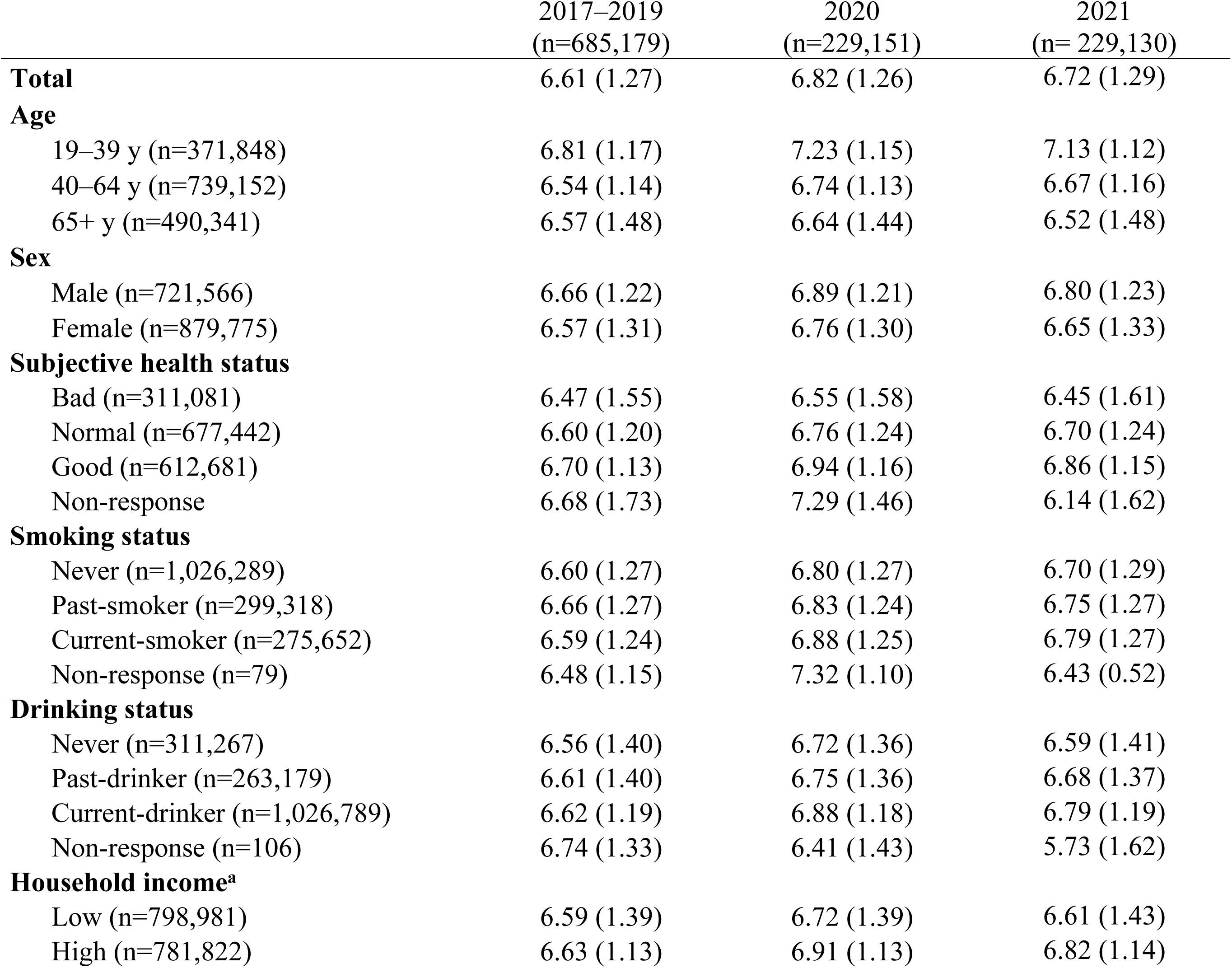
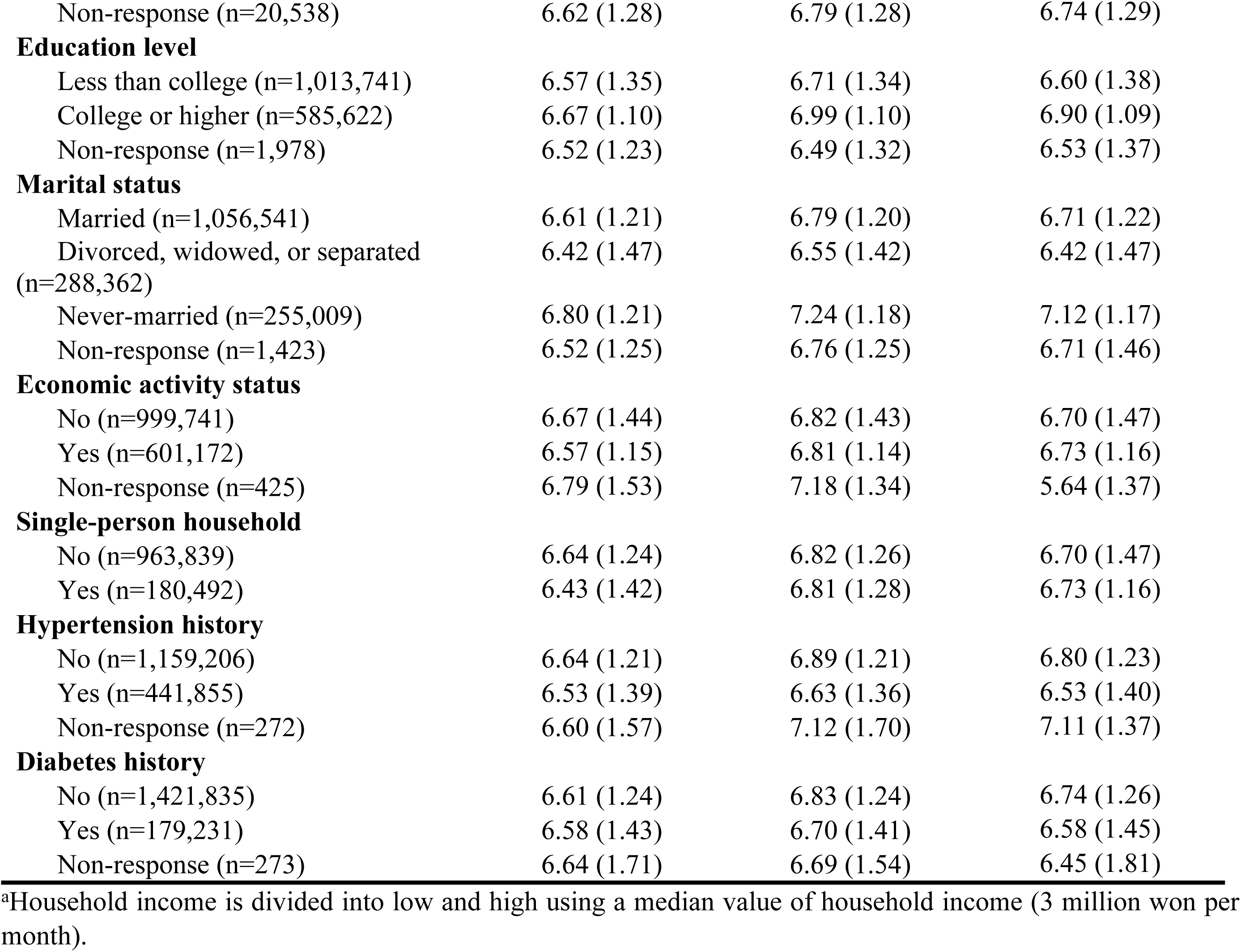
Sleep duration (hr/day) of 1,143,460 study participants; mean (standard deviation)

**Table 2.**
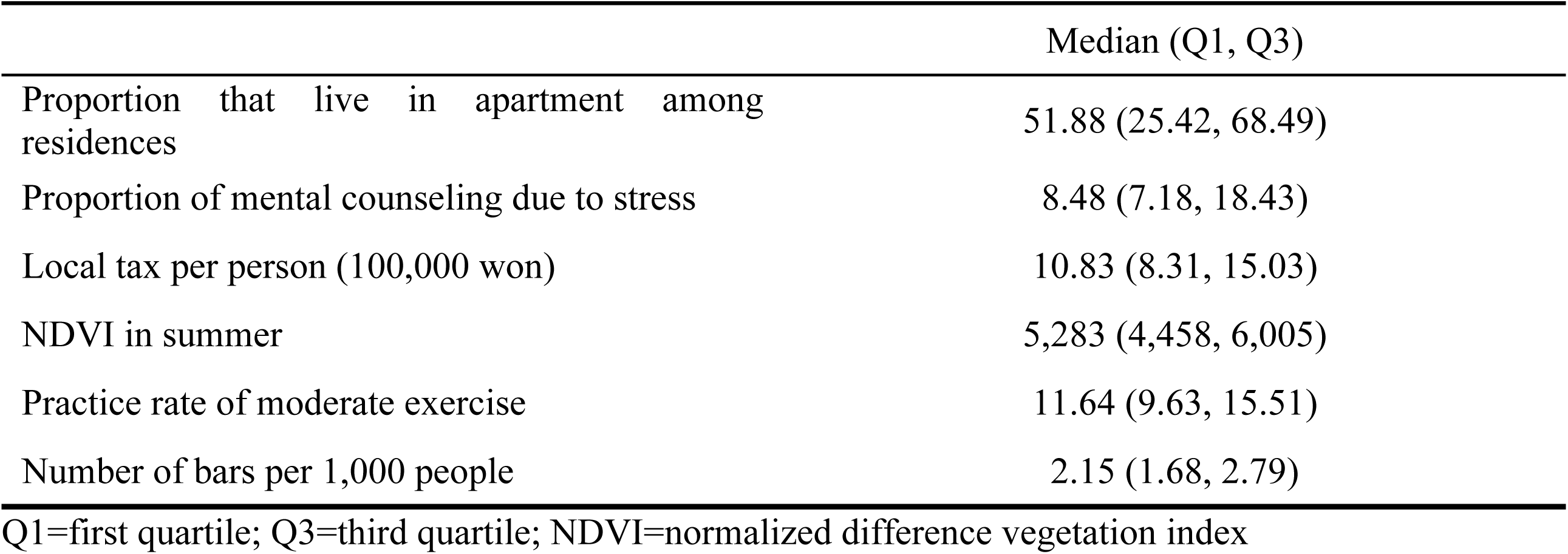
Descriptive statistics for community-level indicators of 229 districts.

|  | Median (Q1, Q3) |
| --- | --- |
| Proportion that live in apartment among residences | 51.88 (25.42, 68.49) |
| Proportion of mental counseling due to stress | 8.48 (7.18, 18.43) |
| Local tax per person (100,000 won) | 10.83 (8.31, 15.03) |
| NDVI in summer | 5,283 (4,458, 6,005) |
| Practice rate of moderate exercise | 11.64 (9.63, 15.51) |
| Number of bars per 1,000 people | 2.15 (1.68, 2.79) |

Q1=first quartile; Q3=third quartile; NDVI=normalized difference vegetation index

Fig. 1 shows the time-trend adjusted change in the sleep duration during the early stage (2020) and long-lasting period (2021) of the pandemic compared to the pre-outbreak period (2017–2019). The daily mean sleep duration increased by 9.66 (95% confidence interval [CI]: 8.53, 10.80) minutes in 2020 and 3.66 (95% CI: 2.09, 5.22) minutes in 2021 compared to that in the 2017–2019 period (Fig. 2). The district-level spatial distribution of the change in sleep duration is displayed in Fig. S1. The increase was more prominent in males, young adults (19– 39 y), employed individuals, those who have a high household income, and highly educated people than the general population (Fig. 3).

**Fig. 1.**
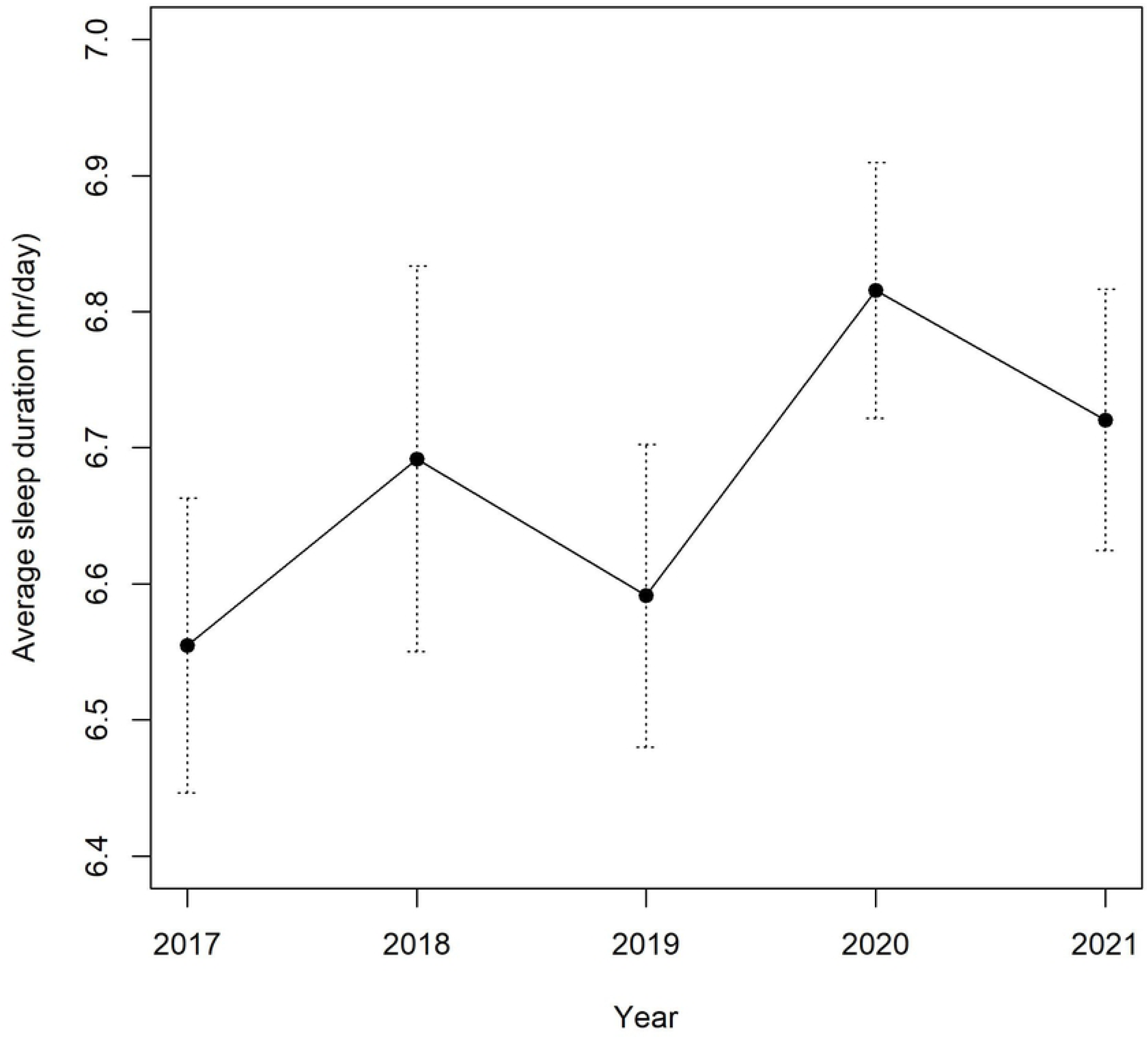
Temporal trend of average sleep duration (min/day) and standard deviation among 229 districts during the study period (2017–2021)

**Fig. 2.**
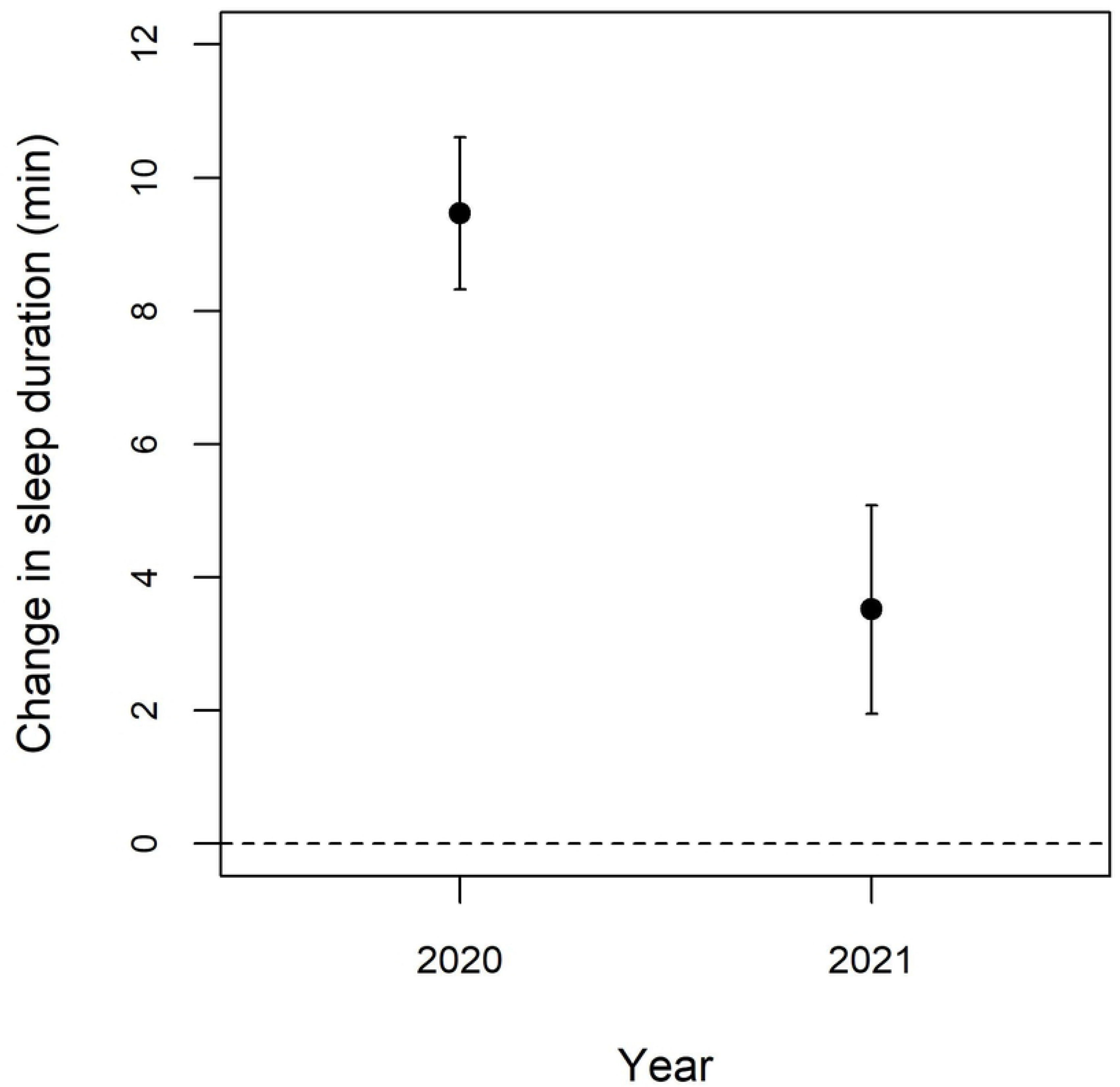
Change in sleep duration during the COVID-19 pandemic (2020 and 2021) compared to the pre-pandemic period (2017–2019)

**Fig. 3.**
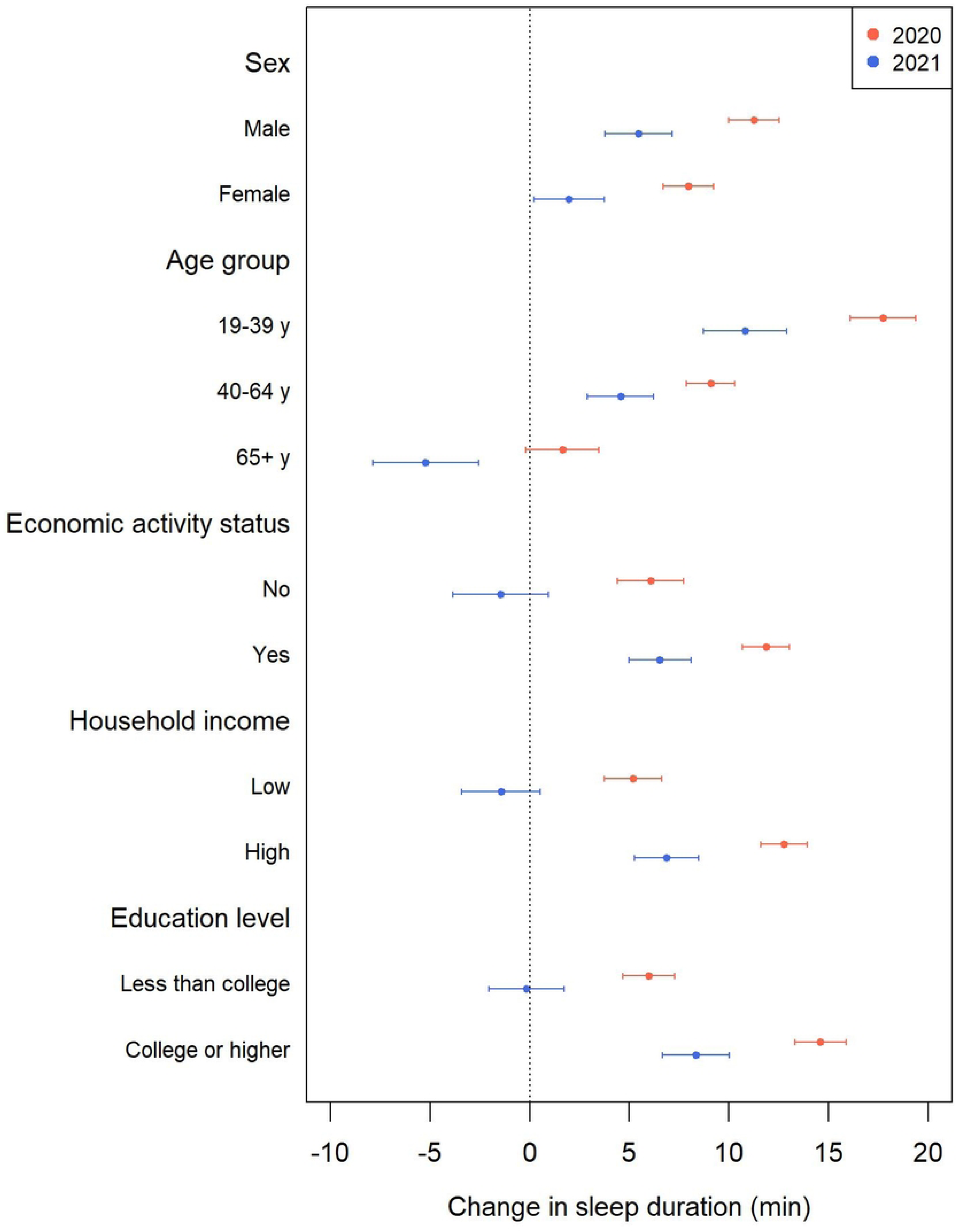
Change in sleep duration during the COVID-19 pandemic (2020 and 2021) compared to the pre-pandemic period (2017–2019) by individual characteristics

We also found an association between the changes in sleep duration during the pandemic and regional characteristics (Table 3). The proportion of apartments demonstrated a positive association with an increase in sleep duration, whereas the NDVI in the summer and the practice rate of moderate exercise had a negative association with the increase in sleep duration in 2020. In 2021, only the practice rate of moderate exercise was associated with an increase in sleep duration. The associations by sex, age, economic activity status, household income, and education level are presented in Tables S1–S5.

**Table 3.** Associations between district-level indicators and changes in sleep duration during (2020 and 2021) and before (2017–2019) the COVID-19 pandemic.

|  | Change in sleep duration<sup>1</sup><br>(95% CI<sup>2</sup>) |  |
| --- | --- | --- |
|  | 2020 | 2021 |
| Proportion of apartments among residences | 2.535 (0.068, 5.001)* | 0.029 (-0.032, 0.090) |
| Proportion of mental counseling due to stress | -0.37 (-1.659, 0.918) | -0.013 (-0.044, 0.019) |
| Local tax per person (100,000 won) | -0.099 (-0.733, 0.535) | 0.004 (-0.012, 0.020) |
| NDVI in summer | -0.002 (-0.003, -0.001)* | 0.000 (0.000, 0.000) |
| Practice rate of moderate exercise | -1.195 (-2.412, 0.022) | -0.032 (-0.062, -0.002)* |
| Number of bars per 1,000 people | -0.482 (-1.310, 0.346) | -0.011 (-0.032, 0.009) |

\*p-value < 0.05
<sup>1</sup>Change in sleep duration was expressed as the change for interquartile range (IQR) increase of the district-level indicators.
<sup>2</sup>CI=confidence interval
Note: These results were calculated by conducting a meta-regression after adjusting for the longitude and latitude of each district.

## Discussion

This study examined the sleep duration during the early stage (2020) and long-lasting period (2021) of the COVID-19 pandemic compared to the pre-outbreak period (2017–2019). We observed that Korean adults increased their sleep duration during the pandemic period, and these changes were associated with the proportion of apartments, NDVI in summer, and the practice rate of moderate exercise. Furthermore, compared to the general population, a higher increase in sleep duration was observed among males, younger adults, those who were employed, and those having higher incomes and education levels.

Our study found that the sleep duration increased by 9.66 minutes in the early stages of the COVID-19 outbreak, which is consistent with previous studies. The average sleep duration increased by 11–19 minutes in the United States and 16 European countries [16], 12– 24 minutes in five metropolitan areas (London, Los Angeles, New York City, Seoul, and Stockholm), and 11 minutes in Seoul, South Korea [20]. Due to the implementation of social restrictions, most individuals spent more time at home and less time on social activities; furthermore, certain workers had to work from home and saved commuting time, which may have led to an increase in sleep duration [26]. At the same time, several psychological symptoms caused by social isolation and pandemic anxiety may also be associated with excessive sleep [11].

Notably, when the COVID-19 pandemic was prolonged in 2021, the sleep duration increased by 3.66 minutes compared to that prior to the outbreak; however, the increase was smaller than that in 2020 when the pandemic began. One study found that sleep patterns slightly returned to pre-pandemic levels from April to June 2020 when the stay-at-home orders were lifted [16], and several studies predicted that changes in sleep patterns would soon return to normal as social distancing and closure policies are eased [19]. Our finding provides empirical evidence for this conjecture, suggesting that people were gradually adapting to the prolonged COVID-19 situation.

In our study, the proportion of apartments was positively associated with changes in sleep duration. This can be explained by the fact that the early stages of the COVID-19 pandemic mainly affected densely populated urban areas in Korea, and accordingly, the social restrictions were implemented more strongly [27]. In contrast, the NDVI in the summer and the practice rate of moderate exercise were negatively associated with an increase in sleep duration. Exposure to green spaces, increase in physical activities, and reduction in time at home can protect against anxiety, fear, and sleep disturbance, and offset the harmful impacts on mental and physical health during the lockdown [28, 29].

We found that sleep duration increased more in males and younger age groups. Because the economic activity rate is higher in males and they spend less time at home on weekday compared to females, the stay-at-home orders may have changed the life pattern of men more significantly than that of women [30, 31]. Furthermore, young individuals who are generally more active in social and economic activities compared to the elderly may have largely reduced their time for activities in response to the social restrictions during the COVID-19 period.

Another finding of our study was a prominent increase in sleep duration in groups with higher socioeconomic status. The effect of the transition of the work process to work-from-home may differ by the state of economic activity, level of education, and household income; households with higher levels of education and income were more likely to get jobs that can be performed at home [32]. Considering that long sleep duration can be negatively associated with mental health outcomes [33, 34], this finding is consistent with a previous study indicating that people with a high socioeconomic status demonstrated a greater increase in mental health problems during the COVID-19 period in South Korea [35].

This study had a few limitations. First, data on sleep duration were collected using a self-reported questionnaire, which can be subject to recall bias. However, as the KCHS was performed by trained interviewers and its quality was systematically managed [22], thus, the bias was expected to be small. Second, because the KCHS is cross-sectional data, and not longitudinal data that follow-up individuals, it is difficult to interpret the results as a causal relationship. Third, our data regarding sleep were limited only to sleep duration, which does not represent the quality of sleep. To closely examine changes in sleep quality during the COVID-19 pandemic, it is necessary to include indicators for sleep quality (e.g., sleep efficiency, sleep latency, and awakenings) in future studies. Despite these limitations, this study has a strength in that it observed changes in sleep duration of the nationwide population during the long-term period of the COVID-19 pandemic. Furthermore, this study revealed that the impact of COVID-19 on sleep duration can be disproportionately distributed by regional and personal characteristics.

## Conclusion

In conclusion, the sleep duration of Korean adults increased during the COVID-19 pandemic, and the increase became smaller as the COVID-19 is prolonged. Importantly, the increase was more noticeable among males, younger adults, employed people, and high socioeconomic groups compared to the general population. Districts with a high proportion of apartments had a greater increase in sleep duration, while districts with a high NDVI in the summer and high moderate exercise rates had a smaller increase in sleep duration during the pandemic. These findings suggest that preventive measures to manage sleep health during a pandemic such as COVID-19 should be framed in consideration of individual and regional characteristics.

## Data Availability

The database of KCHS and community health outcomes and determinants are publicly available from the Korea Centers for Disease Control and Prevention (https://chs.kdca.go.kr/chs/index.do) in Korean. Data of local tax are accessible from the Korea Statistics Office (https://kosis.kr/eng/statisticsList/statisticsListIndex.do?menuId=M_01_01&vwcd=MT_ETITLE&parmTabId=M_01_01#R_18.2). Data of NDVI are available from MODIS (https://modis.gsfc.nasa.gov/data/dataprod/mod13.php).

## Acknowledgements

This work was supported by the Research Program funded by the Korea Centers for Disease Control and Prevention (*fund code: 2733-5788*).

## Authors’ contributions

Conceptualization: JM. Data curation: JM, JO. Formal analysis: JM. Writing—original draft: JM, JO. Writing—review & editing: JM, JO, WL. The author(s) read and approved the final manuscript.

